# Faithful AI in Medicine: A Systematic Review with Large Language Models and Beyond

**DOI:** 10.1101/2023.04.18.23288752

**Authors:** Qianqian Xie, Edward J. Schenck, He S. Yang, Yong Chen, Yifan Peng, Fei Wang

**Affiliations:** Department of Population Health Science, Weill Cornell Medicine, Cornell University; Division of Pulmonary and Critical Care Medicine, New York-Presbyterian Hospital/Weill Cornell Medical Center, 425 E. 61st Street, 4th Floor, Suite 402, New York, NY, USA; Department of Medicine, Weill Cornell Medical College, New York, NY, USA; Department of Pathology and Laboratory Medicine, Weill Cornell Medicine, New York, NY, USA; School of Medicine, University of Pennsylvania

## Abstract

Artificial intelligence (AI), especially the most recent large language models (LLMs), holds great promise in healthcare and medicine, with applications spanning from biological scientific discovery and clinical patient care to public health policymaking. However, AI methods have the critical concern for generating factually incorrect or unfaithful information, posing potential long-term risks, ethical issues, and other serious consequences. This review aims to provide a comprehensive overview of the faithfulness problem in existing research on AI in healthcare and medicine, with a focus on the analysis of the causes of unfaithful results, evaluation metrics, and mitigation methods. We systematically reviewed the recent progress in optimizing the factuality across various generative medical AI methods, including knowledge-grounded LLMs, text-to-text generation, multimodality-to-text generation, and automatic medical fact-checking tasks. We further discussed the challenges and opportunities of ensuring the faithfulness of AI-generated information in these applications. We expect that this review will assist researchers and practitioners in understanding the faithfulness problem in AI-generated information in healthcare and medicine, as well as the recent progress and challenges in related research. Our review can also serve as a guide for researchers and practitioners who are interested in applying AI in medicine and healthcare.

## 1 Introduction

Artificial intelligence (AI) has been gradually applied in different aspects of healthcare and medicine^1–3^. Its benefits are seen in various fields such as quickening medical research, assisting in the detection and diagnosis of diseases, providing tailored health recommendations, and much more (Figure 1). The success of Medical AI has been closely tied to the development of fundamental AI algorithms^4^ and the availability of various biomedical data, including abundant unlabeled data and labeled data annotated by experts.

**Figure 1.**
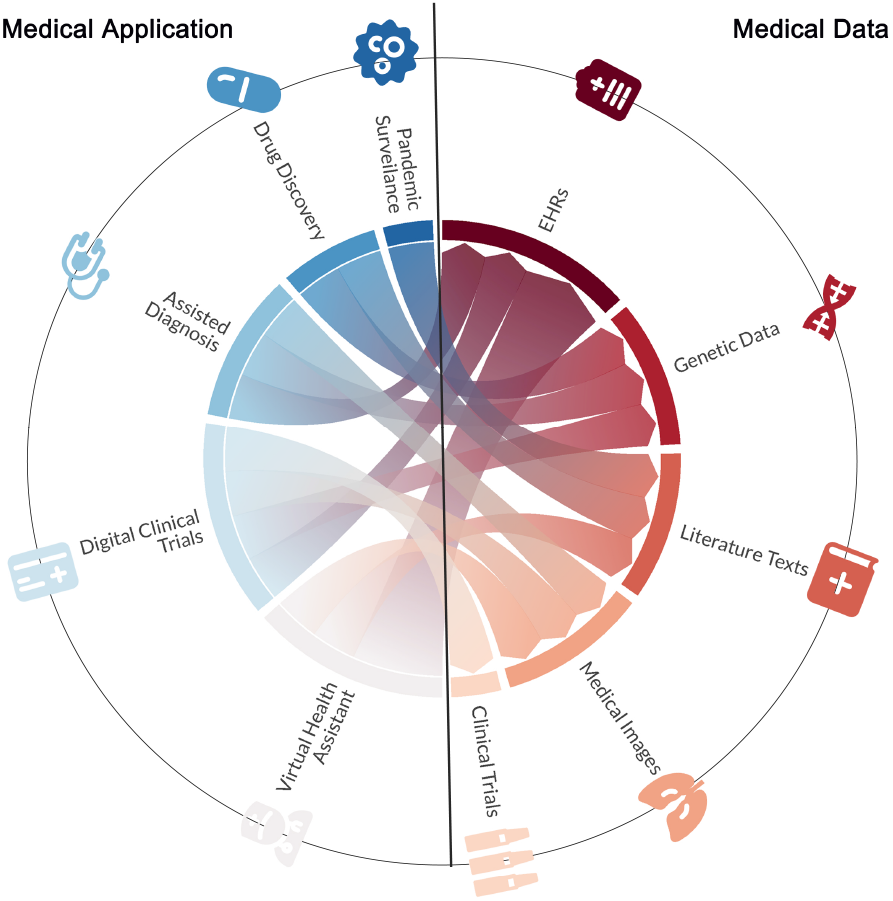
The example of medical data and realistic medical applications with AI.

Recently, pre-trained language models (PLMs)^5^ such as BERT^6^, which are pre-trained on a huge amount of unlabelled language texts in a self-supervised learning manner (typically based on the new backbone neural network architecture called Transformer^7^), have revolutionized natural language processing (NLP). They demonstrated strong potential in Medical AI,5 including tasks beyond NLP like pathology detection^8,9^. More recently, large language models (LLMs)^10^, exemplified by ChatGPT^11^ and GPT-4^12^ developed by OpenAI, have exhibited impressive capabilities to understand complex natural language input and produce human-like text content. LLMs, with their significantly larger model and pre-training data sizes, exhibit remarkably stronger generalizability than PLMs^13^, and unlocked new abilities such as complex reasoning and in-context learning; namely LLMs can perform new tasks without task-specific training but seeing a few examples with task-specific natural language explanation texts^10^. For example, GPT-4 have been reported to pass the threshold of the United States Medical Licensing Exam (USMLE) and showed great potential in assisting clinical decision-making^12,14^. These recent advancements represent a potential turning point for medical AI in several ways: 1) LLMs have remarkable generalization ability and in-context learning ability. This means LLMs can be flexibly and directly applied for a wide range of medical tasks without task-specific training and annotated data. 2) LLMs are designed to be easy to use, accepting natural language as inputs. This makes both laypeople and medical professionals able to access them conveniently. 3) LLMs and fundamental AI techniques are fast-growing. This means they can constantly drive the ongoing development in medical AI.

Despite the promises of medical AI^15^, one major concern gaining considerable attention is its potential risk of generating non-factual or unfaithful information^16^, commonly referred to as the faithfulness problem^17^. Specifically, generative AI methods can generate contents that are factually inaccurate or biased. The faithfulness problem can pose long-term risks, such as preventing medical innovation due to providing wrong guidance in medical research, and misallocating medical resources due to providing unfaithful information on disease treatment. Moreover, ethical issues can be caused by the problem, such as deteriorating trust in healthcare by providing misleading medical information to patients. For example, in Figure 2, a query was prompted to ChatGPT to summarize a systematic review paper named “Diagnostic accuracy of deep learning in medical imaging: a systematic review and meta-analysis” published in npj Digital Medicine 2021^18^. ChatGPT generates the summary with the intrinsic factual error that contradicts the content of the review, such as it showing 82 studies meet the inclusion criteria while the review is actually based on 503 studies. The generated summary also has the extrinsic factual error that can’t be supported by the review, such as “the diagnostic accuracy of deep learning models was higher in computed tomography (CT) and magnetic resonance imaging (MRI) than in radiography and ultrasound, and it was higher in the diagnosis of cancer than in the diagnosis of other diseases”. This could mislead researchers and practitioners and lead to unintended consequences^19^, such as misleading clinical decisions and negative impact on patient care.

**Figure 2.**
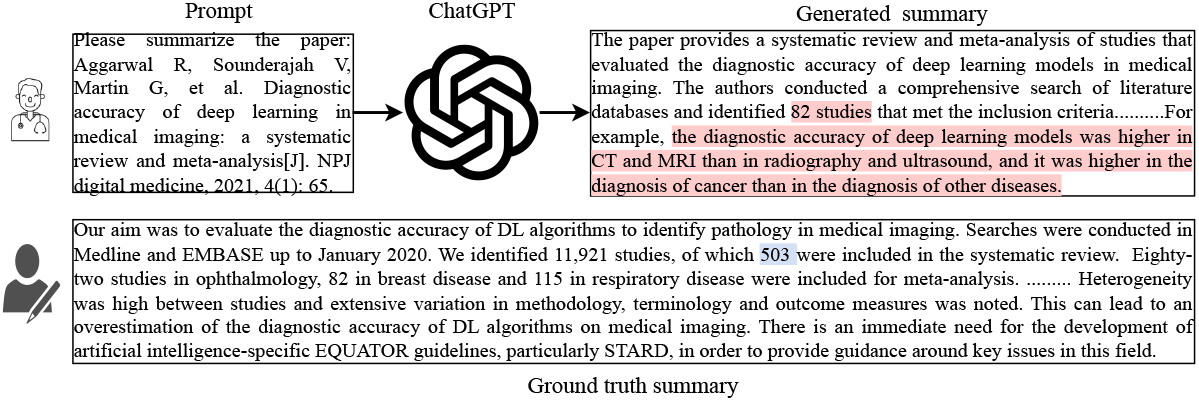
The example of the generated content with factual errors from ChatGPT.

In this review, we provide an overview of the research on the faithfulness problem in existing medical AI studies, including cause analysis, evaluation metrics, and mitigation methods. We comprehensively summarize the recent progress of maintaining factual correctness in various generative medical AI models, including knowledge-grounded large language models, text-to-text generation tasks such as medical text summarization and simplification, multimodality-to-text generation tasks such as radiology report generation, and automatic medical fact-checking. We discuss the challenges of maintaining the faithfulness of AI-generated information in the medical domains, as well as the forthcoming opportunities for developing faithful medical AI methods. The goal of this review is to provide researchers and practitioners with a blueprint of the research progress on the faithfulness problem in medical AI, help them understand the importance and challenges of the faithfulness problem, and offer them guidance for using AI methods in medical practice and future research.

## 2 Search Methodology

### 2.1 Search Strategy

We conducted a comprehensive search for articles published between January 2018 to March 2023 from multiple databases, including PubMed, Scopus, IEEE Xplore, ACM Digital Library, and Google Scholar. We used two groups of search queries:

1) faithful biomedical language models: factuality/faithfulness/hallucination, biomedical/medical/clinical language models, biomedical/medical/clinical knowledge, 2) mitigation methods and evaluation metrics: factuality/faithfulness/hallucination, evaluation metrics, biomedical/medical/clinical summarization, biomedical/medical/clinical text simplification; radiology report summarization, radiology report generation, medical fact-checking.

### 2.2 Filtering Strategy

A total of 3,439 records were retrieved from five databases. 865 articles were kept after removing duplicate records, which were further screened based on title and abstract. We applied the exclusion criteria during the filtering process (also shown in Figure 3): 1) the article is a review paper, 2) the main content of the article is not in English, 3) the article is not related to the medical domain, 4) the article is not relevant to the factuality problem. As a result, 49 articles were retained after the screening process. Then, we conducted a full-text review of these articles and further excluded 12 articles due to the low quality of the content based on the GRADE criteria.

**Figure 3.**
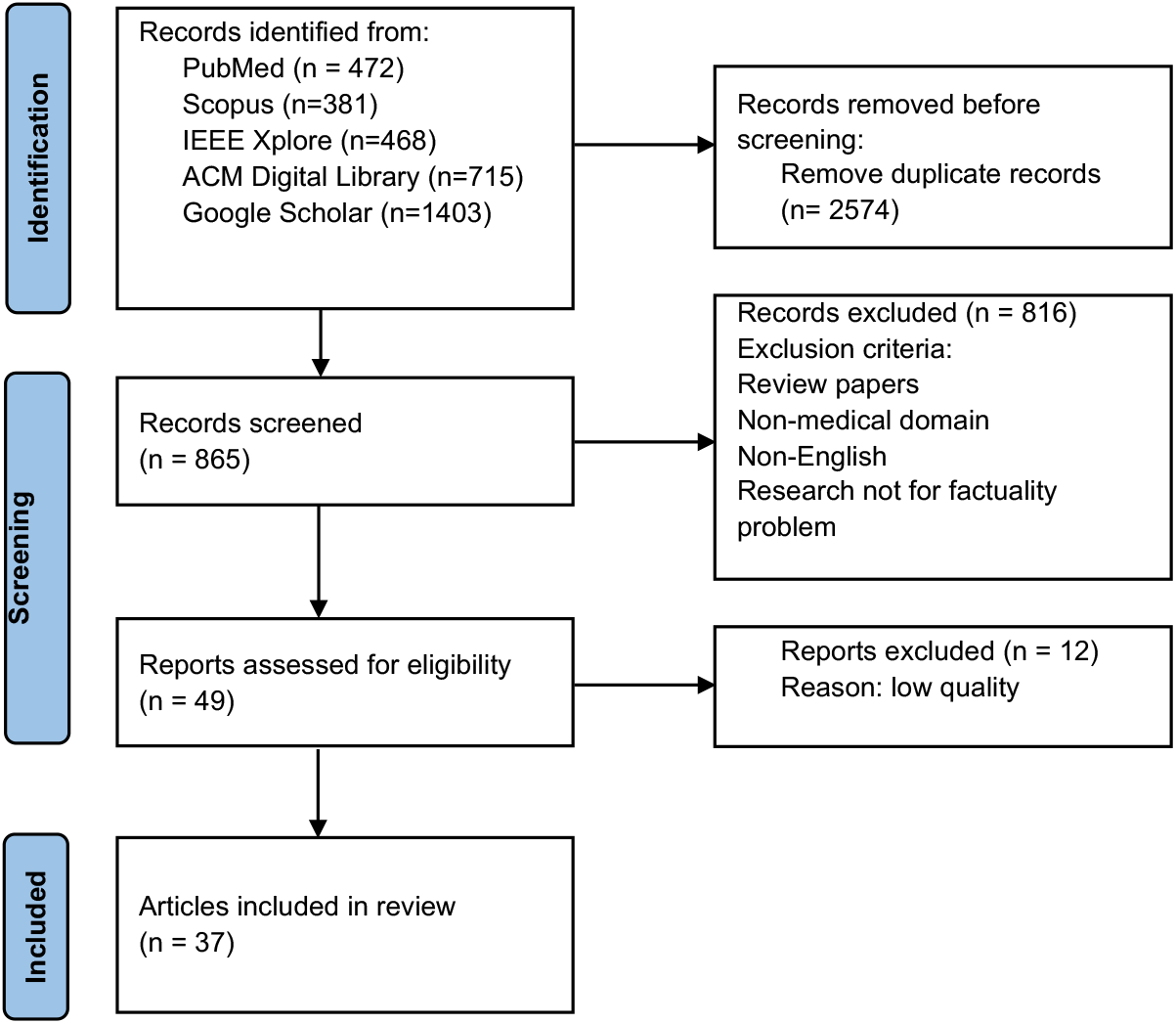
The process of article selection.

## 3 What is Faithfulness Problem ?

### 3.1 Definition and Categorization

In the context of AI or NLP, faithful AI means the algorithm can produce contents that are factually correct, namely staying faithful to facts^17,20^. Generative medical AI systems^21–23^ learn to map from various types of medical data, such as electronic health records (EHRs), medical images, or protein sequences, to desired output, such as the summarization or explanation of medical scans, radiology reports, and three-dimensional (3D) protein structures. A medical AI system is considered unfaithful or to have a factual inconsistency issue, also known as “hallucination” in some studies^20^, if it generates content that is not supported by existing knowledge, reference, or data. Similar to the general generative AI methods, the factual inconsistency issue in generative medical AI generally can also be categorized into the following two major types^24^:

- **Intrinsic Error:** The generated output contradicts with existing knowledge, reference, or data. For example, if there is a sentence in the radiology report summary generated by the AI system, “The left-sided pleural effusion has increased in size”, but the actual sentence in the radiology report is “There is no left pleural effusion”. Then the summary contradicts the facts contained in the input data.
- **Extrinsic Error:** The generated output cannot be confirmed (either supported or contradicted) by existing knowledge, reference, or data. For example, if the content of the radiology report includes: “There is associated right basilar atelectasis/scarring, also stable. Healed right rib fractures are noted. On the left, there is persistent apical pleural thickening and apical scarring. Linear opacities projecting over the lower lobe are also compatible with scarring, unchanged. There is no left pleural effusion”, and the generated summary includes: “Large right pleural effusion is unchanged in size”. Then this piece of information cannot be verified by the given radiology report, since the information about “right pleural effusion” is not mentioned there.

Different from general applications, the tolerance of factual incorrectness should be significantly lower for medical and healthcare tasks, given the high-stakes nature of these fields. Different medical tasks have different references for accessing the factuality of the generated output. For the AI-based text summarization^22^ or simplification systems^25^ that summarize or simplify the input medical texts such as study protocols, biomedical literature, or clinical notes, the reference is usually the input medical document itself, which ensures the AI-generated content is faithful to its original information. For AI systems that automatically generate radiology reports with the input Chest X-ray image^26^, the reference is the radiology report of the input Chest X-ray image written by the radiologist. In table 4, we summarize the reference of different exemplar generative medical tasks for evaluating the factual consistency of the generated output by AI systems.

### 3.2 Why There is the Faithfulness Problem?

The factual inconsistency of medical AI systems can be attributed to a wide range of reasons.

#### 3.2.1 Limitation of backbone language model

Pre-trained language models (PLMs) and the latest large language models (LLMs), commonly used for representing medical data in various types and modalities, have limitations on generalization and representation abilities for medical data. One possible reason is that most existing PLMs and LLMs were not pre-trained with enough medical data, thus, they lack representability there and cannot distinguish between unfaithful information and factually correct information. Although there have been domain-specific PLMs trained with biomedical data such as BioBERT^27^, BlueBERT^28^, PubMedBERT^29^, BioGPT^23^ et al. either with fine-tuning or from scratch, the scale of these data are much smaller than those used for training PLMs in general domains. In addition, most biomedical texts, such as radiology report texts, have limited availability due to privacy concerns. As a result, existing domain-specific language models were mainly pre-trained with biomedical literature texts, that are easy to be accessed on a large scale. Therefore, it is challenging to pre-train LLMs with biomedical domain-specific data that can generalize well and encode comprehensive medical knowledge. This inevitably leads to factual inconsistencies in various medical AI systems that use these PLMs and LLMs as the backbone model.

#### 3.2.2 Data discrepancy

The factual inconsistency problem also arises from the data discrepancy between the ground truth output used for training and the reference in most medical tasks^17,20^. For example, in the medical dialogue generation task, the ground truth output responses are not always faithful to the references, namely dialogue histories, since the topic of responses can be transferred and diverse according to patient input dialogues. Moreover, it is noticed that “ground truth outputs” may be subject to biases and influenced by changing knowledge bases regarding patient-centered goals. Since medical knowledge can evolve and patient needs may shift, the ground truth outputs used in training may not always reflect accurate or up-to-date information. Thus, the medical AI systems that are trained by generating the output with minimized divergence with the ground truth output can produce an output that is not faithful with the reference.

#### 3.2.3 Limitations of decoding

Another cause for the factual inconsistency problem is the limitation of the decoding strategy used by medical AI systems for text generation. There exists the exposure bias problem^30,31^, which is the discrepancy between training and inference on the decoding process based on the decoder. During training, the decoder is encouraged to generate the next token (in the context of NLP, a token means a single unit of meaning in a sentence.) on the conditional of the previous ground truth tokens. While during inference, the ground truth tokens are unobservable, and thus the decoder can only be conditioned on the previously generated tokens by itself to predict the next token. Such discrepancy between training and inference has been shown to potentially result in factual errors of outputs^24^. Moreover, existing methods usually use sampling-based decoding strategies such as beam search and greedy search to improve the diversity of outputs, and they have not considered the factual consistency when selecting tokens and candidate outputs. The randomness of selecting tokens and candidate outputs can increase the probability of generating outputs with factual errors^32^.

## 4 Evaluating and Optimizing Faithfulness in Medical AI

To improve the faithfulness of LLMs, many efforts have focused on improving the backbone model with medical knowledge, optimizing the factual correctness of AI medical systems for various generative medical tasks, and developing fact-checking methods, which will be introduced in the following subsections.

### 4.1 Faithfulness in Large Language Models (LLMs)

Some efforts have focused on explicitly incorporating extra medical knowledge to address the factuality problem in domain-specific PLMs and LLMs. Yuan et al^33^ proposed to train a knowledge-aware language model by infusing entity representations, as well as entity detection and entity linking pre-training tasks based on the Unified Medical Language System (UMLS) knowledge base. The proposed method improves the performance of a series of biomedical language models, such as BioBERT and PubMedBERT, on the named entity recognition and relation extraction tasks. Singhal et al^34^ proposed the Med-PaLM that aligns a 540-billion parameter LLM PaLM^35^ into the medical domains by instruction prompt tuning, which greatly alleviates errors of PaLM on scientific grounding, harm, and bias from low-quality feedback. Their experiments also demonstrate that these LLMs, despite their advancements, are not yet ready to be used in areas such as medicine, where safety is of paramount importance. The instruction prompt tuning used in Med-PaLM proves effective in improving its factuality, consistency, and safety. Zakka et al^36^ proposed the knowledge-grounded language model Almanac, which uses the LLMs as the clinical knowledge base to retrieve and distill information from medical databases for replying to clinical queries, rather than directly generate content with LLMs. Almanac has shown better performance than ChatGPT on medical Q&A based on the 20 questions derived from their ClinicalQA dataset and mitigates the factual inconsistency problem by grounding LLMs with factually correct information retrieved from predefined knowledge repositories. Nori et al^37^ conducted a comprehensive study on the ability of GPT-4 on medical competency examinations and medical Q&A benchmarks recently^34^. On the USMLE self-assessment and sample exam, GPT-4 achieves the average accuracy score of 86.65% and 86.70% and outperforms GPT-3.5 by around 30%. On the multiple-choice medical Q&A benchmark with four datasets, GPT-4 also significantly outperforms GPT-3.5 and the Flan-PaLM 540B^38^ as introduced before by a large margin. Although their assessment highlights the great potential of GPT-4 in assisting healthcare professionals, they suggested GPT-4 still has a large gap in safe adoption in the medical domain like prior LLMs, due to several limitations such as the risk of error generations, biases, and societal issues.

#### Discussion

Although the above studies have shown that the incorporation of medical knowledge into LLMs can potentially help improve their faithfulness. They showed that even SOTA LLMs such as Med-PaLM and GPT-4 cannot satisfy the need for safe use in medicine and healthcare. To bridge the gap, there are several limitations to be addressed and promising future directions.

- Systematic evaluation benchmark: existing methods only assess the factuality of LLMs in limited tasks such as question answering and the United States Medical Licensing Examination (USMLE). There is no systematic evaluation of LLMs in many other critical generative tasks such as medical text generation, and medical applications. We believe future efforts should be spent on evaluating and improving LLMs in diverse medical tasks to fill the gap with domain experts, such as conducting prospective randomized trials to examine the impact of these models on medical diagnosis and treatment.
- Multimodal and multilingual: most existing methods can only process the medical texts and the language in English. Future efforts are encouraged to build LLMs in the medical domains with the ability to tackle inputs with multi-modalities and multiple languages, for example, adapt the recently released multi-modal large language models such as GPT-4 and Kosmos^39^ into the medical domains.
- Unified automatic evaluation method: evaluating the performance of LLMs in factuality is especially challenging in the medical domains and existing methods rely on human evaluation with expert annotation, which is expensive and hard to be on large scale. The unified automatic factuality evaluation method should be proposed to support the effective evaluate the factual correctness of LLMs on various medical tasks.

The above methods only take the initial step, and we consider more efforts should be proposed in the future to make AI methods closer to real-world medical applications.

### 4.2 Faithfulness of AI Models in Different Medical Tasks

Many efforts have been devoted to optimizing the factuality of generative methods in medicine and healthcare, as well as their factuality evaluation for a specific task with various techniques such as incorporating medical knowledge, reinforcement learning, and prompt learning.

#### 4.2.1 Medical Text Summarization

Medical text summarization^40–43^ is an important generative medical task, with the goal of condensing medical texts such as scientific articles, clinical notes, or radiology reports into short summaries. Medical text summarization supports many applications in medicine and healthcare, such as assisting researchers and clinicians to quickly access important information from a large amount of medical literature and patient records and identify key medical evidence for clinical decisions. In the following, we briefly overview the recent efforts in studying the factual inconsistency problem in medical text summarization.

##### Optimization Methods

The factual inconsistency problem was first explored in the radiology report summarization. Specifically, Zhang et al^44^ found that nearly 30% of radiology report summaries generated from the neural sequence-to-sequence models contained factual errors. To deal with the problem, Zhang et al^22^ proposed to optimize the factual correctness of the radiology report summarization methods with reinforcement learning. They evaluated the factual correctness of the generated summary with the CheXpert F1 score^45^, which calculates the overlap of 14 clinical observations between the generated summary and the reference summary. They optimized such factual correctness with policy learning, by taking the factual correctness score of the generated summary as the reward and the summarizer as the agent. They demonstrated that such a training strategy could improve the CheXpert F1 score by 10% when compared with the baseline method. Delbrouck et al^46^ further released the new dataset based on MIMIC-III^47^ with new modalities including MRI and CT, as well as anatomies including chest, head, neck, sinus, spine, abdomen, and pelvis. They proposed the new factual correctness evaluation metric RadGraph score that could be used for various modalities and anatomies and designed a summarization method that optimized the RadGraph score-based reward with reinforcement learning. Their experimental results showed that optimizing the RadGraph score as the reward could consistently improve the factual correctness and quality of the generated summary from the summarizer, where the RadGraph score, F1CheXbert, ROUGE-L^48^ (a commonly used metric in text generation, that calculates the longest common sub-sequence between the generated summary and reference summary) are improved by 2.28%-4.96%, 3.61%-5.1%, and 0.28%-0.5%. Xie et al^49^ proposed the two-stage summarization method FactReranker, which aims to select the best summary from all candidates based on their factual correctness scores. They proposed to incorporate the medical factual knowledge based on the RadGraph Schema to guide the selection of FactReranker. FactReranker achieves the new SOTA on the MIMIC-CXR dataset and improves the RadGraph score, F1CheXbert, and ROUGE-L by 4.84%, 4.75%, and 1.5%.

There are also efforts investigating the factual inconsistency problem in the automatic summarization of other medical texts such as biomedical literature, medical Q&A, and medical dialogues. Deyoung et al^50^ found that the summarizer based on language models such as BART^51^ could produce fluent summaries for medical studies, but the faithfulness problem remains an outstanding challenge. For example, the generated summary from the summarizer only had 54% agree on the direction of the intervention’s effect with that of the input systematic review. Wallace et al^52^ proposed the decoration and sorting strategy that explicitly informed the model of the position of inputs conveying key findings, to improve the factual correctness of the generated summary from the BART-based summarizer for published reports of randomized controlled trials (RCTs). Alambo^53^ studied the factual inconsistency problem of a transformer-based encoder decoder summarization method. They proposed to integrate the biomedical named entities detected in input articles and medical facts retrieved from the biomedical knowledge base to improve the model’s faithfulness. Yadav et al^54^ proposed to improve the factual correctness of the generated summary for medical questions, by maximizing the question type identification reward and question focus recognition reward with the policy gradient approach. Chintagunta et al^55^ investigated using GPT-3 to generate higher-quality labeled data, which has proven to be able to train the summarizer with better factual correctness. Liu et al^56^ proposed the task of automatically generating discharge instructions based on the patients’ electronic health records and the Re3Writer method for the task, which retrieved related information from discharge instructions of previous patients and medical knowledge to generate faithful patient instructions. The human evaluation results showed the patient instructions generated by the proposed method had better faithfulness and comprehensiveness than those generated from baseline sequence-to-sequence methods.

##### Evaluation Metrics

Derivation of effective automatic factual correctness evaluation metrics is critical for evaluating the quality of the generated summary and summarization method development. Existing commonly used evaluation metrics in text summarization, such as ROUGE^48^ and BERTScore^57^, have been proven to be ineffective in evaluating factual correctness, especially in the medical domain. Recently, efforts have been made to develop new metrics for evaluating the factual consistency of the summarizer for different medical texts.

For radiology report summarization, Zhang et al^22^ proposed the CheXpert F1 score that calculates the overlap of 14 clinical observations, such as “enlarged cardiom” and “cardiomegaly”, between the generated summary and the reference summary. Delbrouck et al^46^ further proposed a RadGraph score that calculates the overlap of medical entities and relations based on the RadGraph^58^ (a dataset with annotations of medical entities and relations for radiology reports) between the generated summary and the gold summary, and can be used for various modalities and anatomies. For the biomedical literature summarization, Wallace et al^52^ proposed findings-Jensen-Shannon Distance (JSD) calculating the agreement of evidence directions (including significant difference, or no significant difference) of the generated summary and reference summary of the systematic review, according to JSD. Based on findings-JSD, Deyoung et al^50^ further proposed the improved metric ΔEI that calculates the agreement of the intervention, outcome, and evidence direction based on the Jensen-Shannon Distance, between the generated summary and the input medical studies. Otmakhova et al^59^ proposed the human evaluation approach for medical study summarization, where they defined several quality dimensions, including PICO correctness, evidence direction correctness, and modality to evaluate the factuality of the generated summary. Based on the human evaluation protocol, Otmakhova et al^60^ further developed the Δloss to evaluate the factual correctness of the generated summary on different aspects such as strong claim, no evidence, no claim, etc., and evidence directions. It calculates the difference of negative log-likelihood loss with the summarization model, between the generated summary and the counterfactual summary (the corrupted target summary that has a different modality and polarity from the target summary). Adams et al^61^ did a meta-evaluation on existing automatic evaluation metrics, including BARTScore, BERTScore, CTC^**?**^ and SummaC^62^ (two SOTA metrics based on natural language inference) on assessing long-form hospital-course summarization. Without aligning to the biomedical domain, they found that the evaluation results of these automatic evaluation metrics correlate well with human annotations. However, the calculated correlation is found to has bias and not correct enough, since the generated summaries from LED tend to have a high overlap with original sentences of original inputs rather than regenerate new sentences, which is not applicable in a realistic situation.

##### Discussion

To understand how existing methods perform on the factuality of medical text summarization, in Table 2 and Table 3, we show the performance of SOTA methods on medical study summarization and radiology report summarization with human evaluation and automatic evaluation metrics introduced above. We can see that existing SOTA methods have a relatively good performance on the grammar and lexical of generated summaries for medical studies. Only 8%-9% of the generated summaries are completely factually correct. Less than 50% of the generated summaries are with correct PICO^63^ (Patient, Intervention, Comparison, Outcome) and modality indicating the certainty level of evidence claim (such as strong claim, moderate claim, and weak claim). For radiology report summarization, we can find that although most methods^22,46^ use reinforcement learning for optimizing the factuality, the method proposed in Xie et al^49^ incorporating medical knowledge achieves the best performance, which shows the importance of using medical knowledge to improve factuality. Similarly, the factuality performance of SOTA methods was not high either. For example, the RadGraph F1 scores of SOTA methods were only around 50%.

**Table 1.**
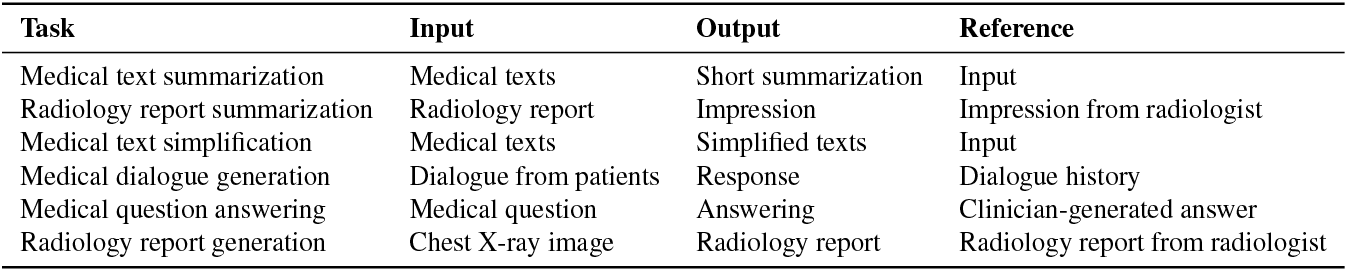
The reference of different exemplar generative medical tasks for evaluating the factual consistency of the generated output by AI systems.

**Table 2.** The performance of human evaluation of SOTA methods on medical study summarization in MS2 dataset^50^ from the paper^59^, which evaluates generated summaries from the factual correctness of PICO, evidence direction, and modality. Factually correct means the composition performance of PICO, evidence direction, and modality.

|  | PICO | Direction | Modality | Factually correct | Grammar | Lexical |
| --- | --- | --- | --- | --- | --- | --- |
| BART<sup>50</sup> | 45% | 77% | 45% | 9% | 75% | 69% |
| LED<sup>50</sup> | 40% | 75% | 44% | 8% | 73% | 73% |

**Table 3.** The performance of SOTA methods for radiology reports summarization on MIMIC-CXR dataset.

|  | ROUGE-1 | ROUGE-2 | ROUGE-L | F1CheXbert | RadGraph |
| --- | --- | --- | --- | --- | --- |
| L4-RL<sup>46</sup> | 51.96 | 35.65 | 47.10 | 74.86 | 48.23 |
| FactReranker<sup>49</sup> | 55.94 | 40.63 | 51.85 | 76.36 | 53.17 |

**Table 4.** Summary of datasets.

| Data | Domain | Claim Source | Negation Method | Size |
| --- | --- | --- | --- | --- |
| PUBHEALTH<sup>80</sup> | Public health | Fact checking and News websites | Natural | 11,832 |
| HEALTHVER<sup>83</sup> | COVID | TREC-COVID<sup>88</sup> | Natural | 14,330 |
| SCI-FACT<sup>81</sup> | Biomed | S2ORC<sup>89</sup> | Human | 1,409 |
| COVID-Fact<sup>84</sup> | COVID | Reddit | Automatic | 4,086 |
| Misinformation<sup>86</sup> | Medical | Fact checking and News websites | Human | 573 |
| Covert<sup>85</sup> | COVID | Twitter | Human | 300 |
| RedHOT<sup>87</sup> | Health | Reddit | Human | 22,000 |

For future work, we believe the following aspects are important to focus on.

- *Creation of benchmark data sets*. There are limited open source datasets^5^ for medical text summarization, due to the high cost of expert annotation and other issues such as privacy. In fact, there are only two public datasets for medical study^50,52^ and radiology report summarization, which covers limited modalities, anatomies and languages. Moreover, the sizes of these datasets are much smaller when compared with datasets in the general domain. High-quality large-scale benchmark datasets should be created in the future to facilitate the development of medical summarization methods.
- *Derivation of unified automatic evaluation metrics*. Existing evaluation metrics for assessing the factuality of methods in different medical texts are specific, and there are even no such metrics for methods on certain types of medical texts, such as medical dialogue and medical question summarization. It is important to develop unified automatic evaluation metrics for supporting the assessment of summarization methods across different types of medical text.
- *Development of better optimization methods*. Most existing methods employ reinforcement learning to improve the factuality of generated summaries, while little attention has been paid to incorporating medical knowledge. As mentioned before, the factual correctness of SOTA methods is not good enough for reliable use in realistic medicine and healthcare. Therefore, more advanced methods are needed, for example, by using more powerful backbone language models and designing a more effective decoding strategy guided by medical facts.

#### 4.2.2 Medical Text Simplification

Medical text simplification^64^ aims to simplify highly technical medical texts to plain texts that are easier to understand by non-experts such as patients. It can greatly improve the accessibility of medical information.

##### Optimization Methods

Lu et al^65^ proposed the summarize-then-simplify method for paragraph-level medical text simplification, where they designed the narrative prompt with key phrases to encourage the factual consistency between the input and the output. The human evaluation showed that their proposed method significantly outperforms the BART-based simplification method proposed in^25^ by 0.49 on the 5-point scale on the factuality of outputs. Jeblick et al^66^ utilized ChatGPT to simplify 45 radiology reports and asked 15 radiologists to evaluate outputs from factual correctness, completeness, and potential harm. They found the radiologists agreed that most of the outputs were factually correct, but there were still some errors, such as misinterpretation of medical terms, imprecise language, and extrinsic factual errors. Lyu et al^67^ also evaluated the performance of ChatGPT on simplifying radiology reports to plain language. The evaluation dataset consisted of 62 chest CT screening reports and 76 brain MRI screening reports. ChatGPT showed good performance with an overall score of 4.268 in the five-point system assessed by radiologists. It has an average of 0.08 and 0.07 places of information missing (assessing the number of places with information lost) and inaccurate information (assessing the number of places with inaccurate information). The suggestions for patients and healthcare providers generated by ChatGPT tended to be general, such as closely observing any symptoms, and only 37% provided specific suggestions based on the findings of reports. ChatGPT also showed instability in generating over-simplified reports, which can be alleviated by designing better prompts. Overall, the factuality problem in automatic medical text simplification methods has rarely been explored, and we believe more efforts should be devoted to this area.

##### Evaluation Metrics

Similar to the medical text summarization, automatic similarity-based evaluation metrics such as ROUGE and BERTScore were also used for evaluating the semantic similarity between outputs and references in the medical text simplification. Moreover, other important aspects in the evaluation are the readability and simplicity of outputs, for which commonly used metrics include the Flesch-Kincaid grade level (FKGL)^68^, the automated readability index (ARI)^69^, and SARI^70^. Intuitively, the language model pre-trained on the technical corpus can assign higher likelihoods to the technical terms than the language models pre-trained on the general corpus. Based on this intuition, Devaraj et al.^25^ proposed a new readability evaluation metric calculating the likelihood scores of input texts with a masked language model trained on the technical corpus. Devaraj et al^71^ further proposed a method for evaluating factuality in text simplification based on RoBERTa to classify factual errors, including insertion, deletion, and substitution errors based on human-annotated data. However, they found that metrics such as ROUGE and their proposed method must be revised to effectively capture factual errors.

##### Discussion

We can see that there is limited research studying the factuality problem in medical text simplification. Moreover, the assessment of the factuality of existing methods has been largely relying on human evaluation. Therefore, it is important to have more efforts to study the factuality problem in medical text simplification by developing better optimization methods, automatic evaluation metrics, and creating more data resources in the future.

#### 4.2.3 Radiology Report Generation

Radiology report generation aims to automatically generate radiology reports illustrating clinical observations and findings with the input medical images such as chest X-rays and MRI scans. It can help to reduce the workload of radiologists and improve the quality of healthcare.

##### Optimization Methods

Most existing efforts adopted reinforcement learning (RL) to optimize the factual correctness of radiology report generation methods. Nishino et al^72^ proposed the RL-based method by optimizing the clinical reconstruction score, calculating the correctness of predicting finding labels with the generated report, to improve the factual correctness of generated reports. Their experiments showed that the model improved the performance on the F1 score of the factuality metric CheXpert (it assesses the clinical correctness of the generated report that calculates the overlap of finding labels between generated and reference report) by 5.4% compared with the model without the RL-based optimization. Miura^26^ proposed to use reinforcement learning to optimize the entailing entity match reward assessing the inferentially consistent (such as entailment, neutral, contradiction) between entities of the generated report and the reference report, and exact entity match reward evaluating the consistent of disease and anatomical entities between the generated report and reference report, which encourages the model to generate key medical entities that are consistent with references. The proposed method greatly improves the F1 CheXpert score by 22.1% compared with the baselines. However, it relied on the named entity recognition methods that are not trained with annotated data of the Chest X-ray domain. Delbrouck et al^73^ further designed the RadGraph reward calculating the overlap of entities and relations between the generated report and the reference, based on the RadGraph dataset including annotated entities and relations of the Chest X-ray reports. The proposed method improves the factuality evaluation metric F1 RadGraph score (calculating the overlap of entities and relations between the generated report and the reference) by 5.5% on the MIMIC-CXR dataset when compared with baselines including^26^. Nishino et al^74^ further proposed an RL-based method of Coordinated Planning (CoPlan) with the fact-based evaluator and description-order-based evaluator, to encourage the model to generate radiology reports that are not only factually consistent but also chronologically consistent with reference reports. Their method outperforms the baseline T5 model on clinical factual accuracy by 9.1%.

##### Evaluation Metrics

Similar to medical text summarization, to evaluate the clinical correctness of the generated radiology reports, some efforts^26,72,75^ proposed to use the CheXpert-based metric to evaluate the overlap of 14 clinical observations between generated reports and references annotated by the CheXpert. Delbrouck et al^73^ proposed the RadGraph-based metric to calculate the overlap of the clinical entities and relations between generated reports and references annotated by the RadGraph schema. Recently, Yu et al^76^ examined the correlation between existing automatic evaluation metrics including BLEU^77^, BERTScore, F1 CheXpert, and RadGraph F1, and the score given by radiologists on evaluating the factuality of the generated reports. They found that the evaluation results of F1 CheXpert and BLEU were not aligned with that of radiologists, and BERTScore and RadGraph F1 were more reliable. They further proposed a new evaluation metric, RadCliQ, which is the weighted sum of the score from BLEU and RadGraph F1 based on their optimized coefficients. It showed better alignment with the evaluation of radiologists than the above four metrics.

##### Discussion

Existing methods have demonstrated the effectiveness of using RL and medical knowledge in improving the factuality of generated radiology reports. However, the medical knowledge is typically incorporated in implicit ways based on RL, we consider that future efforts should pay more attention to explicitly incorporating medical knowledge on improving the encoder and decoder of the LLMs. For example, investigating the use of knowledge grounded backbone language models as encoder^34^, and developing decoding strategy guided by medical facts^49,78^. Moreover, the radiology report generation task requires a combination of information, both radiology images and the associated text reports. We believe cross-modality vision-language foundation models^79^ should be explored to improve the faithfulness of radiology report generation methods in the future. For the evaluation metrics, there is only one paper^76^ as we described above on analyzing the correlation between automatic factuality evaluation metrics and scores of experts based on human annotation. It is necessary to have more effort on developing automatic factuality evaluation metrics and creating public benchmark datasets to help the meta-evaluation of these metrics.

### 4.3 Medical Fact Checking

Automatic medical fact-checking aims to detect whether the claim made in certain medical texts is true, which is a promising method for assisting in detecting factual errors and improving the factuality of medical generative methods. Many existing efforts have contributed to the creation of medical data resources to facilitate the development of automatic medical fact-checking methods and their evaluation. Kotonya et al.^80^ built the PUBHEALTH dataset with 11.8K public health-related claims with gold-standard fact-checking explanations by journalists. Wadden et al.^81^ created the SCI-FACT dataset with 1.4K clinical medicine-related scientific claims paired with abstracts including their corresponding evidence, and the annotated labels (including supports, refutes, and neutral) as well as rationales. Poliak et al^82^ collected over 2.1K verified question-answer pairs related to COVID-19 from over 40 trusted websites. Sarrouti et al^83^ developed HEALTHVER for evidence-based fact-checking of health-related claims, with 14,330 claim-evidence pairs from online questions on COVID-19, and their label including supports, refutes, and neutral. On a relevant effort, Saakyan et al^84^ created COVID-Fact with 4,086 real-world claims related to the COVID-19 pandemic, sentence-level evidence for these claims, and their counterclaims. Mohr et al^85^ built a fact-checked corpus with 300 COVID-19-related tweets annotated with their verdict label, biomedical named entities, and supporting evidence. Srba et al^86^ developed a new medical misinformation dataset with 573 manually and more than 51k automatically annotated relations between verified claims and 317k news articles, including the claim presence label indicating whether a claim is included in the article and article stance label indicating whether a claim is supported by the article such as supports, refutes, and neutral. Wadhwa et al^87^ constructed the RedHOT corpus with 22,000 health-related social media posts from Reddit annotated with claims, questions, and personal experiences, which can support identifying health claims and retrieving related medical literature.

There were also efforts to develop automatic medical fact-checking methods with these data resources. Kotonya et al^80^ proposed an explainable automatic fact-checking method with a classifier for predicting the label of the given clain based on pre-trained language models such as BERT, SciBERT^90^, BioBERT, and a summarization model based on the BERTSum summarizer^91^ for generating fact-checking explanations. On the PUBHEALTH dataset, the SciBERT-based prediction method achieved the highest macro F1, precision, and accuracy scores, and fact-checking explanation model fine-tuned on the PUBHEALTH dataset achieved promising performance. Wadden et al^81^ proposed the automatic fact-checking pipeline with the SCI-FACT dataset that retrieves abstracts based on input claims according to the TD-IDF similarity, selects rationale sentences and then predicts the labels (SUPPORTS, REFUTES, or NOINFO) of abstracts regarding the given claims with BERT based related language models. They investigated using SciBERT, BioMedRoBERTa, RoBERTa-base, and RoBERTa-large as the sentence encoder, where the RoBERTa-large achieves the best performance on label prediction. Wadden et al^92^ proposed MULTIVERS for predicting the fact-checking label for a claim and the given evidence abstract, which uses Long-former encoder^93^ to encode the long sequence from the claim and the evidence abstract and predicts the abstract-level fact-checking label aligned to the sentence-level label by multi-task learning. On three medical fact-checking datasets, including HEALTHVER, COVID-Fact, and SCI-FACT, MULTIVERS showed better performance on the zero-shot and few-shot settings compared with existing methods, due to the weak supervision by the multi-task learning.

#### Discussion

Although fact-checking is a promising way of detecting and mitigating the hallucination problem of AI methods, we can see that existing fact-checked datasets only covered limited medical facts in specific domains such as COVID and public health. Therefore, future efforts can also be spent on developing effective automatic fact-checking methods based on other resources with rich medical facts, such as medical knowledge bases and plain medical texts. Moreover, there are no methods exploring fact-checking on LLMs, which should be a research focus as well, given the popularity of LLMs.

## 5 Limitations and Future Directions

With all the reviews and discussions of existing works on faithful AI in healthcare and medicine above, we will summarize the overall limitations of existing studies and discuss future research directions.

### 5.1 Datasets

#### Unlabeled Data for Self-supervised Learning

PLMs and LLMs trained with self-supervised learning, requiring large-scale unlabeled medical data. However, collecting such data is difficult due to privacy and cost considerations. For example, the unlabeled clinical data used to train the clinical PLMs such as ClinicalBERT^94^ is 3·7GB, while that used to train LLMs such as GPT-4 can be up to 45TB. Moreover, most datasets are limited to a single language, where English is predominantly used, and single data modality. This can hinder the development of faithful medical AI methods in low-resources and rural areas. Therefore, it’s crucial to develop multimodal, multilingual PLMs to improve the generalization and faithfulness of medical language models, using various languages and data types, such as text and images.

#### Annotated Data for Supervised Learning and Evaluation

Developing and evaluating faithful medical AI methods relies on high-quality annotated medical data. Collecting large-scale, high-quality annotated medical data is even more challenging due to the high cost of both time and expertise. Furthermore, existing annotated datasets are usually small, with no publicly accessible data for the meta-evaluation of automated metrics in most medical tasks. This makes it difficult to verify the reliability of these metrics. Thus, building domain expert annotated datasets for various medical tasks is essential to analyze metric alignment with expert preferences and develop reliable automatic metrics and effective mitigation methods.

### 5.2 Backbone Models

#### Biomedical Domain-Specific Language Models

Many existing medical AI methods use biomedical PLMs and fine-tune them with task-specific datasets for various downstream tasks. However, these models, typically pre-trained with the biomedical literature texts and a few other types of medical texts, capture limited medical knowledge. Moreover, their sizes are typically small (usually less than 1B parameters). For example, up to now, the largest PLMs in the medical domain, PubMedGPT, has 2·7B parameters, which is far smaller than the scale of LLMs in the general domain (e.g., GPT-4 with 100T parameters). Therefore, to improve the reliability of biomedical PLMs, future strategies could include larger model sizes and multimodal data training.

#### Large Generative Language Models

LLMs have shown amazing natural language understanding and generation abilities. However, they are not mainly trained with data in the medical domain, and none are publicly available, hindering the development of reliable medical AI. Therefore, adapting LLMs for the medical field is vital, with strategies like fine-tuning with domain-specific data and prompt tuning with human feedback. There is a recent work^34^ that aligns the LLM PaLM with the medical domain. Unfortunately, it is still not publicly available.

### 5.3 Faithful Medical AI Methodologies

#### Mitigation Methods

Although factuality is a critical issue in existing medical AI methods, little effort has been devoted to improving the faithfulness of backbone language models and medical AI methods for downstream tasks. No research has investigated the factuality in medical tasks, including medical dialogue generation, medical question answering, and drug discovery et al. It is also important to develop explainable medical AI methods, especially LLMs. The explanation can play an important role in alleviating the hallucination problem. It helps to understand and trace causes of factual errors, makes it easier to assess factual errors, and enhances the medical AI faithfulness.

#### Incorporating Medical Knowledge

Existing efforts have proven the effectiveness and importance of improving the factuality in both backbone language models and medical AI methods for specific tasks by incorporating medical knowledge. However, most focused on extracting medical knowledge from external biomedical knowledge bases. Recently, there has been an effort21 investigating the efficiency of instruction prompt tuning on injecting medical knowledge into LLMs, which relies on human feedback and thus can be expensive and time-consuming. Effective incorporation of medical knowledge in efficient and scalable ways remains a critical challenge.

### 5.4 Evaluations

#### Automatic Evaluation Metrics

Existing automatic evaluation metrics, calculating the overlap of medical facts between outputs generated by the algorithm and references, fail to distinguish different types of factual errors, such as intrinsic and extrinsic errors, as introduced in Section 3.1. In addition, the assessment of factuality relies on human evaluations for many tasks, such as medical question answering, without automatic factuality evaluation metrics. Future work should explore fine-grained automatic metrics to clarify and assess varied medical factual errors and a unified evaluation guideline for standardized criteria across medical tasks.

#### Meta-evaluation

Assessing the effectiveness of automatic evaluation metrics is critical for correctly evaluating the factuality of methods. Otherwise, the ineffective automatic evaluation metrics can misguide the optimization and evaluation of methods. There is rare work^76^ investigating the meta-evaluation of automatic factuality metrics used in various medical tasks and analyzing their alignment with domain experts. Therefore, it is important to conduct the prospective randomized controlled trial (RCT) level assessment for these evaluation metrics to evaluate their reliability and effectiveness.

## 6 Conclusions

The advancement of fundamental AI methods, particularly the latest LLMs, provides great opportunities for medical AI. However, significant concerns exist about the reliability, safety, and factuality of the content generated by medical AI methods. This review provides the first comprehensive overview of the faithfulness problem in medical AI by analyzing its causes, summarizing mitigation methods and evaluation metrics, discussing challenges and limitations, and envisioning future directions. Existing research on investigating the factuality problem in medical AI is still in its early stages, and there are substantial challenges to data resources, foundation models, mitigation methods, and evaluation metrics in this area. Future research efforts should focus on tackling these challenges and exploring opportunities for novel, faithful medical AI research that involve adapting LLMs, prompt learning, and other techniques. We hope this review will inspire further investigation in this direction and serve as a guide for researchers and practitioners to safely implement AI methods in real-world medical practice. By encouraging the development of more reliable and accurate medical AI systems, we can contribute to improved patient care and better clinical decision-making.

## Data Availability

All data produced in the present study are available upon reasonable request to the authors

## Author Contributions

QQX, Conceptualization, Literature search, Figures, Writing - original draft. QQX played a key role in conceptualizing the paper, managing the collection and organization of relevant literature, and drafting the original manuscript. EJS, Conceptualization, Writing - review & editing. EJS was involved in the conceptualization, reviewed, and edited the manuscript. HSY, Conceptualization, Writing - review & editing. HSY was involved in the conceptualization, reviewed, and edited the manuscript. YFP, Writing - review & editing. YFP reviewed and edited the manuscript. YC, Writing - review & editing. YC reviewed and edited the manuscript. FW, Conceptualization, Supervision, Funding Acquisition, Project Administration, Writing - review & editing. FW reviewed the manuscript, provided overall supervision of the project, was instrumental in acquiring funding for the research, and administrated the entire project.

## Data Availability

The datasets are available from authors upon reasonable request.

## Acknowledgement

The work was supported by NSF 1750326, NIH R01AG080624, R01AG076448, R01AG080991, R01AG076234, and RF1AG072449.

